# Cerebral Venous Sinus Thrombosis Associated with SARS-CoV-2; a Multinational Case Series

**DOI:** 10.1101/2020.09.12.20186106

**Authors:** Ashkan Mowla, Banafsheh Shakibajahromi, Shima Shahjouei, Afshin Borhani-Haghighi, Nasrin Rahimian, Humain Baharvahdat, Soheil Naderi, Fariborz Khorvash, Davar Altafi, Seyed Amir Ebrahimzadeh, Ghasem Farahmand, Alaleh Vaghefi Far, Vijay K. Sharma, Saeideh Aghayari Sheikh Neshin, Georgios Tsivgoulis, Ramin Zand

## Abstract

**Background:** SARS-CoV-2 induced coagulopathy can lead to thrombotic complications such as stroke. Cerebral venous sinus thrombosis (CVST) is a less common type of stroke which might be triggered by COVID-19. We present a series of CVST cases with SARS-CoV-2 infection.

**Methods:** In a multinational retrospective study, we collected all cases of CVST in SARS-CoV-2 infected patients admitted to nine tertiary stroke centers from the beginning of the pandemic to June 30^th^, 2020. We compared the demographics, clinical and radiological characteristics, risk factors, and outcome of these patients with a control group of non-SARS-CoV-2 infected CVST patients in the same seasonal period of the years 2012-2016 from the country where the majority of cases were recruited.

**Results:** A total of 13 patients fulfilled the inclusion criteria (62% women, mean age 50.9± 11.2 years). Six patients were discharged with good outcomes (mRS≤2) and three patients died in hospital. Compared to the control group, the SARS-CoV-2 infected patients were significantly older (50.9 versus 36.7 years, *p*<0.001), had a lower rate of identified CVST risk factors (23.1% versus 84.2%, *p*<0.001), had more frequent cortical vein involvement (38.5% versus 10.5%, *p*: 0.025), and a non-significant higher rate of in-hospital mortality (23.1% versus 5.3%, *p*: 0.073).

**Conclusion:** CVST should be considered as potential comorbidity in SARS-CoV-2 infected patients presenting with neurological symptoms. Our data suggest that compared to non-SARS-CoV-2 infected patients, CVST occurs in older patients, with lower rates of known CVST risk factors and might lead to a poorer outcome in the SARS-CoV-2 infected group.

## 1. Introduction

Although strokes due to cerebral venous sinus thrombosis (CVST) are much less common than arterial strokes, it afflicts younger patients and its mortality is still high in developing countries.(1) There is a long list of predisposing factors for CVST but post-viral CVST is a less known entity.(2)

Coronavirus disease 19 (COVID-19) caused by severe acute respiratory syndrome coronavirus 2 (SARS-CoV-2), is a rapidly growing pandemic with significant impacts on global health. The growing knowledge of COVID-19 shows multiple systems involvement.(3)

Coagulopathy is a feature of SARS-CoV-2 infection. Disseminated intravascular coagulation, elevated D-dimer, fibrinogen level, fibrin/fibrinogen degradation product, thrombocytopenia, and the presence of antiphospholipid antibodies are associated with the COVID-19 and its severity.(4, 5) Hypercoagulable states can lead to complications of COVID-19 including venous thromboembolic events, myocardial infarctions, and stroke.(6, 7) Several studies have discussed the rate and characteristics of arterial stroke in the setting of SARS-CoV-2 infection.(7-9) However, only a few case reports have described CVST associated with SARS-CoV-2.(10-14) Furthermore, we are not aware of any study that compared the characteristics of SARS-CoV-2 infection associated CVST with non-SARS-CoV-2 infected CVST.

Herein, we present a series of cases of CVST in patients positive for SARS-CoV-2 who were admitted to tertiary stroke centers and compared their characteristics with non-SARS-CoV-2 infected CVST patients from the country where the majority of cases were recruited.

## 2. Materials and Methods

### 2.1. Study design and patients

This multicenter observational study was conducted and reported according to the Strengthening the Reporting of Observational Studies in Epidemiology (STROBE),(15) and Enhancing the QUAlity and Transparency Of health Research (EQUATOR) guidelines.(16) We included adult hospitalized patients with a definite diagnosis of CVST and confirmed diagnosis of SARS-CoV-2 infection based on reverse transcription-polymerase chain reaction (RT-PCR) for SARS-CoV-2 or typical COVID-19 symptoms and corresponding findings on chest computed tomography (CT) where RT-PCR was not available. In each collaborating center, a board-certified radiologist with advanced neuroradiology training confirmed the diagnosis of CVST according to the following criteria: detecting a cord-like filling defect in a cerebral venous sinus site on brain Magnetic Resonance Venogram (MRV) or CT Venogram (CTV), an absence of a flow void with alteration of signal intensity in the dural sinus on Magnetic Resonance Imaging (MRI), a cord-like hyperdensity on non-enhanced CT in any of the cerebral venous sinus locations.(17) Cortical vein thrombosis was diagnosed based on the presence of a cord- or dot-like hyperintensity/ hyperdensity on conventional MRI or non-contrast CT in a major cortical vein site or detecting filling defects in the superficial cortical veins on CTV or MRV.(18) We attributed the CVST to SARS-CoV-2 infection if the infection symptom onset or diagnosis (whichever was earlier) occurred between two weeks prior to the onset of CVST symptoms and two days after hospital admission for CVST. CVST was not attributed to SARS-CoV-2 infection in the cases who initially did not have a positive test and then developed the symptoms of SARS-CoV-2 infection beyond two days after CVST diagnosis due to the possibility of nosocomial infection.

### 2.2. Data collection

The core investigators invited their existing networks and collaborators from several countries through a personal phone call and email conversations. The investigators and collaborators further introduced the study to local societies for physicians, academic health systems and hospitals that were designated as COVID-19 centers, sometimes using local languages. We received CVST cases attributed to the SARS-CoV-2 infection from the beginning of pandemic to June 30^th^, 2020 from nine tertiary university hospitals in Iran, Singapore, and the United States. Many other centers in Europe and Asia were also contacted; however, they did not have cases or were not able to meet our timeline.

Data for each patient were collected on a standardized electronic data gathering form. Data were gathered from patients’ medical records, images on picture archiving and communication systems, and hospital information systems. Demographic data, clinical presentation, COVID-19 severity, timelines of both diseases, radiological findings including the involved sinuses, the presence of cortical vein thrombosis and hemorrhagic or ischemic parenchymal lesions, CVST risk factors, admission laboratory findings, management, outcome based on modified Rankin Scale (mRS) and in-hospital mortality were assessed. We considered intraparenchymal hemorrhagic conversion of venous infarct in the setting of CVST as intraparenchymal hemorrhage (IPH). The severity of the SARS-CoV-2 infection was assessed at the onset of the CVST presentation. We asked the collaborators to classify the severity of SARS-CoV-2 infection in the reported cases according to the world health organization guidelines(19) into the following categories: asymptomatic, mild, moderate, severe, and critical. Both clinical and chest radiographic findings were considered to determine the severity of the disease.

The interval between the onset of SARS-CoV-2 infection and CVST was calculated by subtracting the date of COVID-19 onset of symptoms or diagnosis (whichever was earlier) from the date of onset of CVST presentation.

For the control group, we used a previously collected dataset of 57 CVST patients who were admitted between February 20 and June 30 in the years 2012 to 2016 from the country where the majority of cases were recruited, Iran. This dataset was driven from a previous retrospective cohort study.(20) We intended to match the cohorts in terms of seasonal variation since the CVST characteristics and risk factors may follow a seasonal pattern.(21) The studied variables and their definitions, the units, and the methods of observation were reviewed in the control group to ascertain that they are consistent between the cases and the control group.

The study protocol was designed by the investigators at the Neuroscience Institute of Geisinger Health System, Pennsylvania, USA, and received approval by the Institutional Review Board (IRB) of Geisinger Health System and other participating institutions when it was required.

### 2.3. Outcome measures

The primary study outcome measure was disability at discharge, as assessed by means of the modified Rankin scale, a global measure of disability on a seven-level scale, with scores ranging from 0 (no symptoms) to 6 (death).(22) We considered mRS≤2 as good outcome. The in-hospital mortality was regarded as death due to any cause in the same hospital admission. The same outcome measures were used previously in the control group.

### 2.4. Statistical analysis

We reported mean± standard deviation (SD) for quantitative variables and frequencies and relative frequencies for qualitative variables. We compared the demographic, clinical, and radiological characteristics and the outcome between the current series of patients and those described in a previous cohort from the same country where most cases were recruited from with independent sample T-test, Chi-Square and Fisher’s exact test. We did not perform any further regression analysis due to our limited sample size.(23) A P-value of less than 0.05 was considered significant. We applied SPSS software for statistical analysis.(24)

## 3. Results

We received 13 eligible cases from nine centers in three countries. The mean age was 50.9± 11.2 years, ranging from 32 to 71 years. Eight patients (61.5%) were women.

The diagnosis of SARS-CoV-2 infection was made by chest CT and typical symptoms in one patient and by RT-PCR in the others. In two patients, CVST was diagnosed with brain CTV and in the others by MRV. In the initial brain imaging, four patients had IPH, and two presented with venous infarcts. Initial head CT without contrast of two of these patients are shown in Figure 1. The others had no parenchymal lesions on the initial brain imaging.

**Figure 1.**
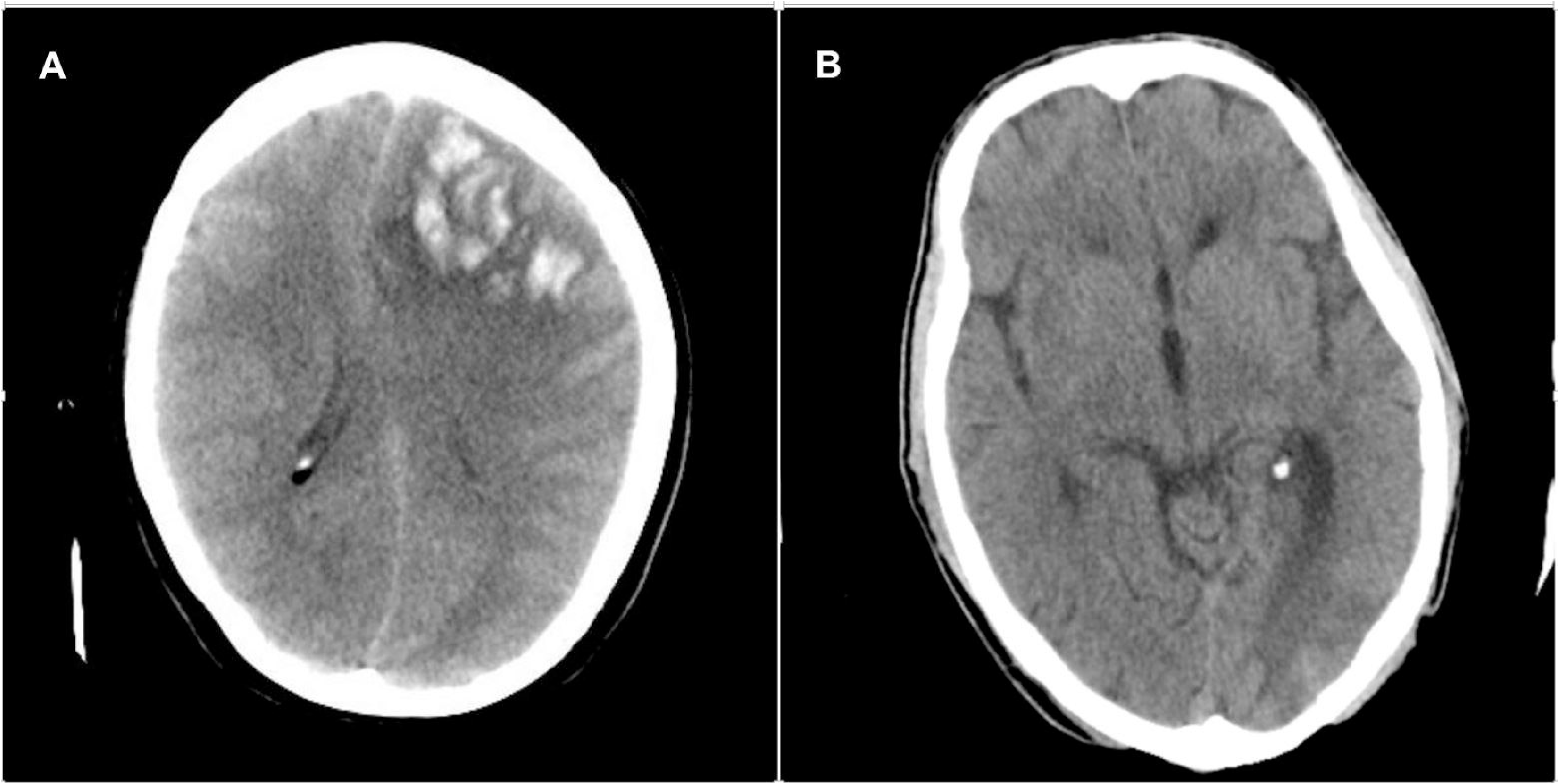
A: Initial Head CT scan without contrast (axial section) of a middle aged woman with superior sagittal sinus thrombosis shows venous infarct of the left anterior frontal lobe with hemorrhagic conversion along with mass effect and midline shift. B: Initial Head CT scan without contrast (axial section) of a middle aged woman with left transverse and sigmoid sinus thrombosis shows a venous infarct in the left occipital lobe.

Demographic data, clinical presentation, radiological findings, and outcome of the patients are presented and compared with the non-SARS-CoV-2 infected CVST cohort in Table 1. Compared to the control group, CVST patients positive for SARS-CoV-2 infection were significantly older (50.9± 11.2 years versus 36.7± 12.7 years, *p*< 0.001). In only three SARS-CoV-2 infected patients (23.1%), a known risk factor for CVST, oral contraceptive use, was identified, whereas, in the control group, 48 patients (84.2%) had at least one identified risk factor (*p*<0.001). Cortical vein involvement was significantly more frequent among patients with SARS-CoV-2 infection (five patients (38.5%) versus six patients (10.5%), *p:* 0.025). Furthermore, the in-hospital mortality rate was higher among SARS-CoV-2 infected patients; however, the difference did not reach statistical significance (23.1% vs. 5.3%, *p*: 0.073).

**Table 1.**
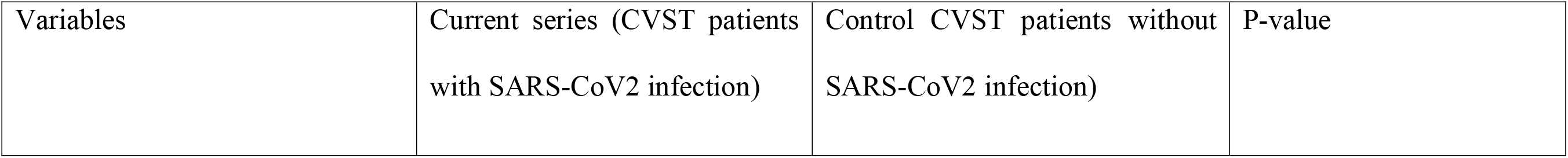
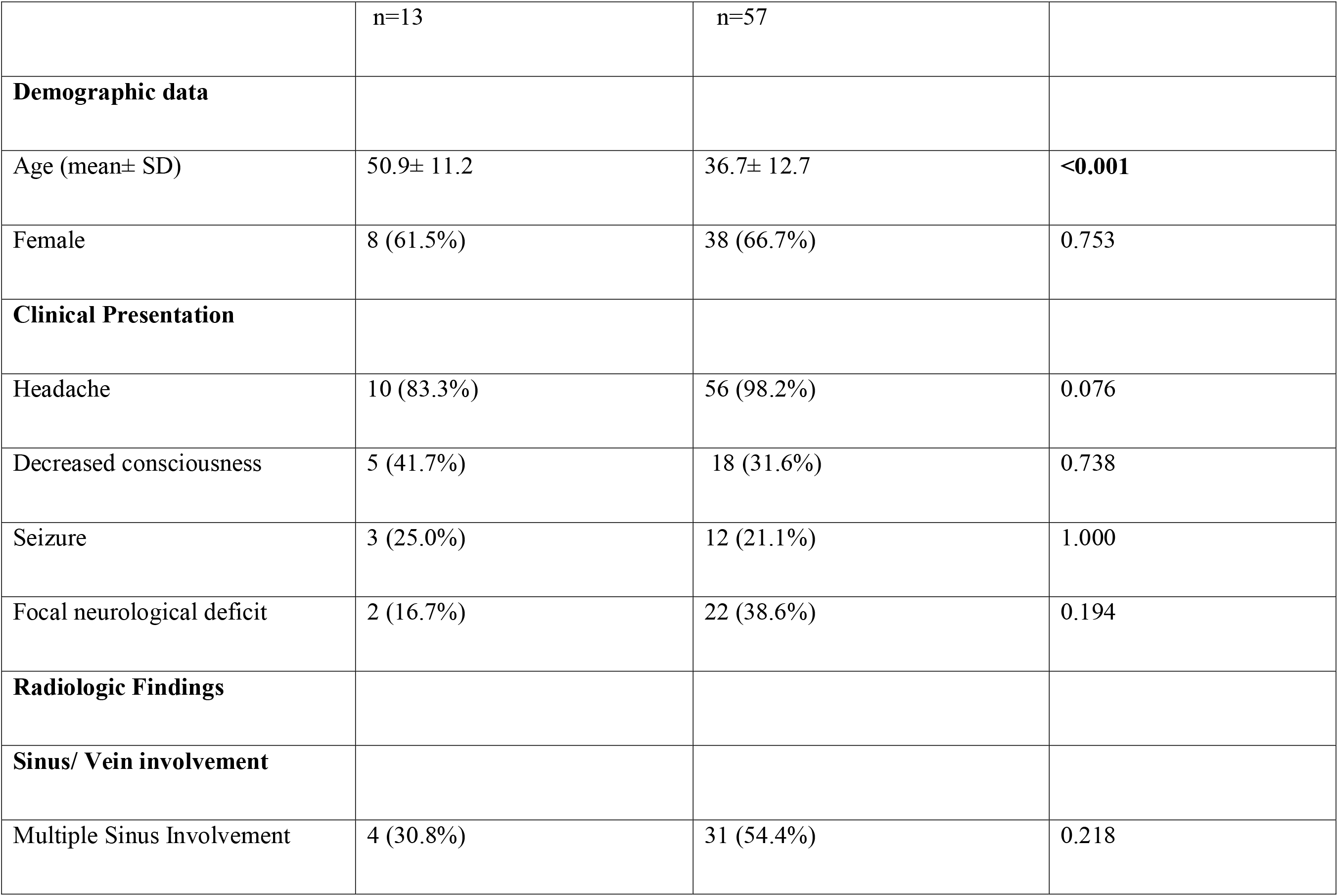
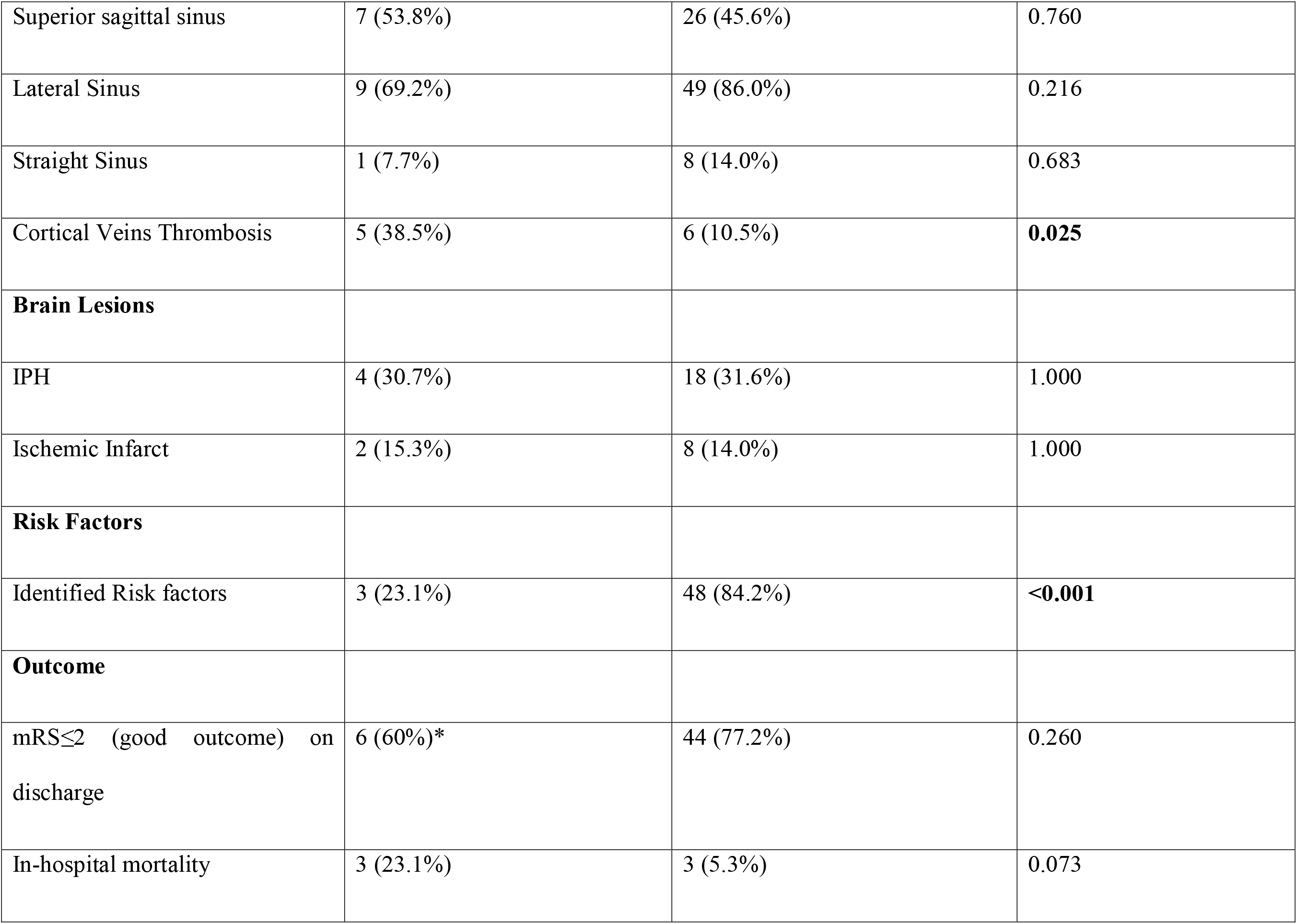
Comparison of demographic, clinical and radiological characteristics and outcome of patients with SARS-CoV-2 infection with those without SARS-CoV-2 infection. **IPH**= Intraparenchymal Hemorrhage, **mRS**= modified Rankin Scale *Frequency is calculated among ten patients with available mRS scores at discharge.

One patient had no COVID-19 associated symptoms. Nine patients had mild to moderate respiratory symptoms and one presented with severe COVID-19 associated symptoms. The COVID-19 severity data for two patients were not available. In four patients, the SARS-CoV-2 infection was detected on the same day of CVST presentation. The remaining patients presented with COVID-19-associated symptoms prior to CVST symptom onset. In none of the patients, symptoms, signs, or imaging findings suggestive of extracranial venous thromboemboli including deep vein thrombosis or pulmonary emboli was reported. Table 2 demonstrates the baseline laboratory data of these patients.

**Table 2.**
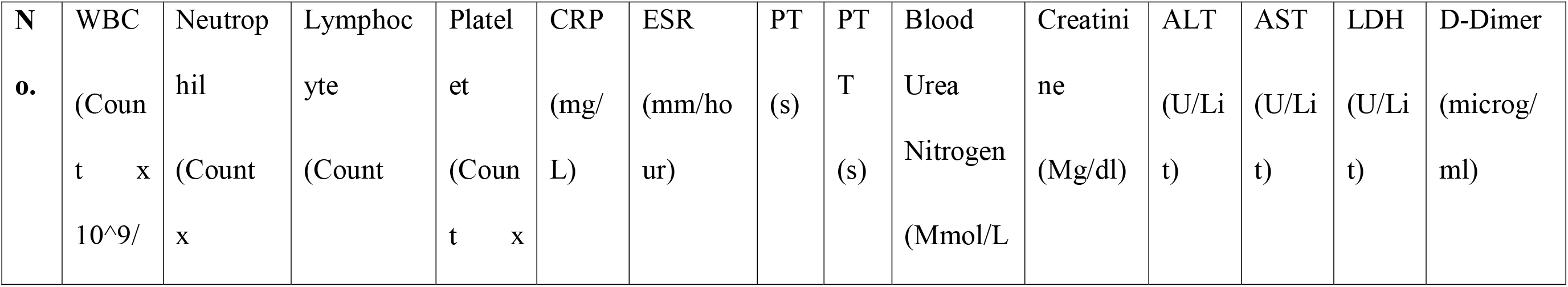
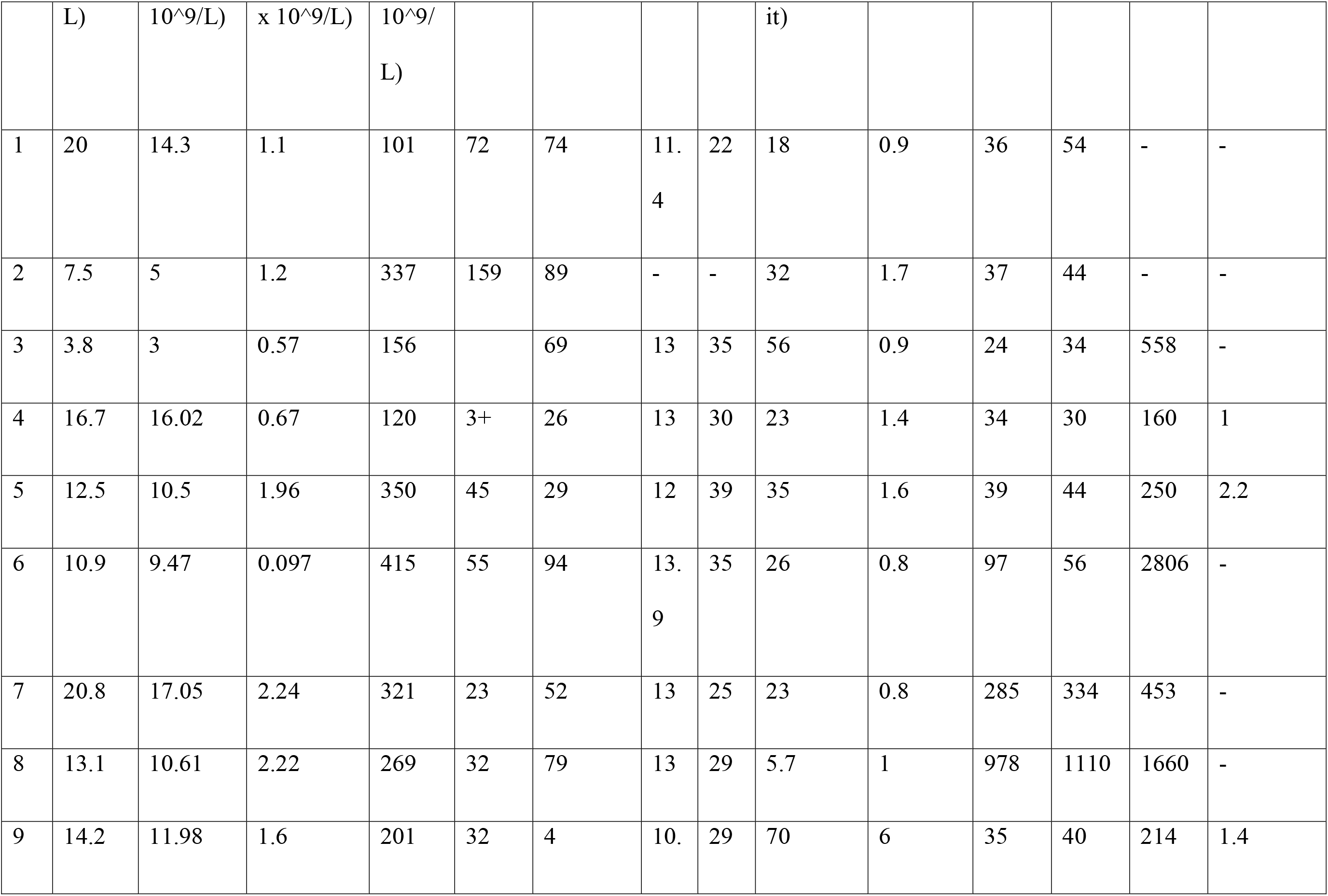
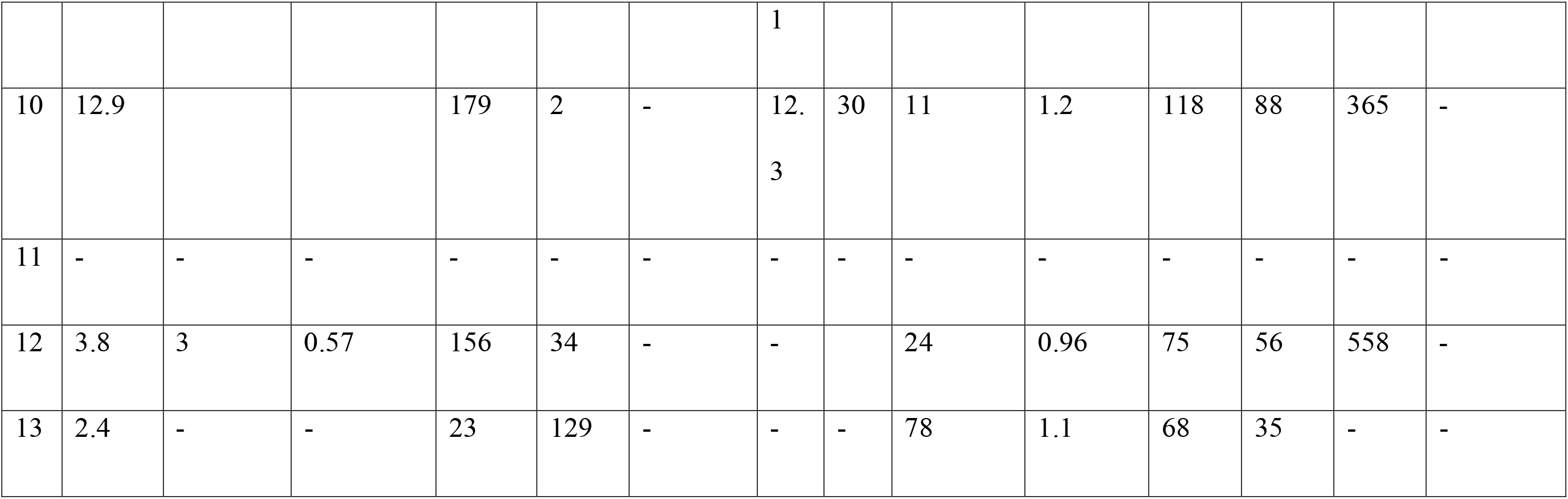
Baseline laboratory data of patients with CVST and COVID-19 **WBC:** White blood cell count, **CRP**: C - reactive protein, **ESR**: Erythrocyte sedimentation rate, **PT**: Prothrombin time, **PTT**: Partial Thromboplastin time, **ALT**: Alanine Aminotransferase, **AST**: Aspartate Aminotransferase, **LDH**: Lactate Dehydrogenase

All patients received anticoagulant therapy and one underwent decompressive hemicraniectomy. The mean length of hospital stay was 18.7 ± 13.1 days. Six patients (60% of the patients with available mRS scores at discharge) were discharged with good outcomes (mRS≤2) and three patients died in hospital. One patient was still hospitalized at the time of data analysis and mRS score at discharge were not available for three patients.

## 4. Discussion

We reported a case series of 13 patients who were simultaneously diagnosed with the SARS-CoV-2 infection and CVST from Iran, Singapore and the United States. The mean age was significantly higher in the SARS-CoV-2 positive patients compared to the CVST patients in the control cohort. The development of CVST in SARS-CoV-2 infected patients with older age also has been reported in several recent cases.(11, 13, 14) Our patients had a significantly lower rate of identified known CVST risk factors. Higher age and significantly less identified risk factors for CVST among our patients compared to the non-SARS-CoV-2 infected control group suggest that SARS-CoV-2 infection might be the triggering factor for CVST.

The mechanisms through which SARS-CoV-2 can trigger CVST are not fully understood. Cytokine release and systemic inflammation, endothelial cell injury, and prothrombotic effects of unopposed angiotensin II can lead to a hypercoagulable state in COVID-19.(25) Sepsis-induced dehydration, blood stasis due to immobility in such patients, and direct neuroinvasion might be the additional mechanisms.(26, 27)

In this study, one patient was asymptomatic and most patients had mild to moderate COVID-19 symptoms at the onset of CVST. Our data indicated that the severity of SARS-CoV-2 infection respiratory symptoms might not necessarily be associated with the risk of thrombotic cerebral complications such as CVST. This finding is in concordance with the previous reports of cerebrovascular events in the patients with COVID-19.(7, 10, 13, 28) Since the asymptomatic or mild cases of SARS-CoV-2 infection might not receive the diagnosis prior to the CVST presentation, it is suggested to screen for SARS-CoV-2 in CVST cases during the current pandemic, particularly in those with no other CVST risk factors to avoid the late diagnosis and subsequent nosocomial spread of the infection.

Headache was the most frequent symptom of our cases and is the main presenting symptom of CVST in up to 95% of patients. (20) Decreased level of consciousness was another major symptom of CVST in this case series. Headache and decreased consciousness have been reported in 13% and 7.5% of COVID-19 patients, respectively.(29) As such, CVST should be considered as potential comorbidity in COVID-19 patients who present with headache and decreased consciousness, even in those with mild COVID-19 symptoms.

Hemorrhagic lesions (IPH) were detected in the initial brain imaging in four patients (30.7%). Brain hemorrhagic lesions have been found in up to 35% of CVST patients.(30) Thus, in SARS-CoV-2 infected patients who present with brain hemorrhagic lesions, CVST should be considered as a potential underlying cause.

Although it was not statistically significant, our analysis suggested that the SARS-CoV-2-attributed CVST cases had poorer outcomes with higher in-hospital mortality compared with the non-SARS-CoV-2 infected CVST patients. Systemic Inflammation, respiratory compromise and multi-organ damage in the setting of CVST can contribute to the poorer outcome seen in this series. Furthermore, cortical vein thrombosis was significantly more frequent in our cases compared to the non-SARS-CoV-2 infected CVST patients. Concomitant cortical vein thrombosis could be associated with poor outcome in CVST patients.(18) In our case series, all three patients who died in the hospital course had cortical vein involvement in addition to sinus thrombosis.

To the best of our knowledge, our study is the largest case series to date on patients with SARS-CoV-2 infection who developed CVST; however, it has several limitations. It has a small size; however, given it is the largest case series on this topic up to the present time, it can still provide valuable information about such patients compared with the non-SARS-CoV-2 infected patients. Additionally, there might be a degree of selection bias. Since our cases were received from particular tertiary centers that were invited to participate, and the majority of the patients were recruited from one country, they might not properly represent the population of COVID-19 patients who present with neurological symptoms; nevertheless, we selected the control cohort from the same country. Furthermore, there were several missing data in a couple of our patients, and given it is a multicenter study, laboratory data were not unified in all centers. In addition, the lack of a complete inherited thrombophilia assessment in some cases could be a source of bias in the interpretation of our analysis on identified CVST risk factors.

## 5. Conclusion

In conclusion, our case series study showed that SARS-CoV-2 might trigger CVST. Health care providers should consider CVST as potential comorbidity in the SARS-CoV-2 infected patients who present with the neurological symptoms. Our data suggest that compared to non-SARS-CoV-2 infected patients, CVST occurs in older patients with a lower rate of known CVST risk factors and might lead to a poorer outcome in SARS-CoV-2 infected patients.

## Data Availability

The data that support the findings of this study are available from the corresponding author upon reasonable request.

## Acknowledgments

We would like to thank Dr. Elahe Mohammadi Vosough and Dr. Hamid Reza Mirkarimi for their invaluable contribution to our data collection.

